# SARS-CoV-2 Humoral Immune Responses in Convalescent Individuals Over 12 Months Reveal Severity-Dependent Antibody Dynamics

**DOI:** 10.1101/2023.12.05.23299462

**Authors:** Nadia Siles, Maisey Schuler, Cole Maguire, Dzifa Amengor, Annalee Nguyen, Rebecca Wilen, Jacob Rogers, Sam Bazzi, Blaine Caslin, Christopher DiPasquale, Melissa Abigania, Eric Olson, Janelle Creaturo, Kerin Hurley, Todd A. Triplett, Justin F. Rousseau, Stephen M. Strakowski, Dennis Wylie, Jennifer Maynard, Lauren I. R. Ehrlich, Esther Melamed

**Affiliations:** Department of Neurology, Dell Medical School, University of Texas at Austin, Austin, Texas; Department of Molecular Biosciences, University of Texas at Austin, Austin, Texas; Department of Chemical Engineering, University of Texas at Austin, Austin, Texas; Babson Diagnostics, Austin, Texas; Department of Neurology, Dell Medical School, University of Texas at Austin, Austin, Texas; Dell Seton Medical Center at the University of Texas, Austin, Texas; Department of Oncology Dell Medical School, University of Texas at Austin, Austin, Texas; Department of Immunotherapeutics & Biotechnology, School of Pharmacy, Texas Tech University Health Sciences Center, Abilene, TX, USA; Department of Neurology, Dell Medical School, University of Texas at Austin, Austin, Texas; Department of Population Health, Dell Medical School, University of Texas at Austin, Austin, Texas; Department of Psychiatry, Indiana University School of Medicine, Indianapolis; Department of Psychiatry, Dell Medical School, University of Texas at Austin, Austin, Texas; Center for Biomedical Research Support, University of Texas at Austin, Austin Texas

## Abstract

**Background:** Understanding the kinetics and longevity of antibody responses to SARS-CoV-2 is critical to informing strategies toward reducing Coronavirus disease 2019 (COVID-19) reinfections, and improving vaccination and therapy approaches.

**Methods:** We evaluated antibody titers against SARS-CoV-2 nucleocapsid (N), spike (S), and receptor binding domain (RBD) of spike in 98 convalescent participants who experienced asymptomatic, mild, moderate or severe COVID-19 disease and in 17 non-vaccinated, non-infected controls, using four different antibody assays. Participants were sampled longitudinally at 1, 3, 6, and 12 months post-SARS-CoV-2 positive PCR test.

**Findings:** Increasing acute COVID-19 disease severity correlated with higher anti-N and anti-RBD antibody titers throughout 12 months post-infection. Anti-N and anti-RBD titers declined over time in all participants, with the exception of increased anti-RBD titers post-vaccination, and the decay rates were faster in hospitalized compared to non-hospitalized participants. <50% of participants retained anti-N titers above control levels at 12 months, with non-hospitalized participants falling below control levels sooner. Nearly all hospitalized and non-hospitalized participants maintained anti-RBD titers above controls for up to 12 months, suggesting longevity of protection against severe reinfections. Nonetheless, by 6 months, few participants retained >50% of their 1-month anti-N or anti-RBD titers. Vaccine-induced increases in anti-RBD titers were greater in non-hospitalized relative to hospitalized participants. Early convalescent antibody titers correlated with age, but no association was observed between Post-Acute Sequelae of SARS-CoV-2 infection (PASC) status or acute steroid treatment and convalescent antibody titers.

**Interpretation:** Hospitalized participants developed higher anti-SARS-CoV-2 antibody titers relative to non-hospitalized participants, a difference that persisted throughout 12 months, despite the faster decline in titers in hospitalized participants. In both groups, while anti-N titers fell below control levels for at least half of the participants, anti-RBD titers remained above control levels for almost all participants over 12 months, demonstrating generation of long-lived antibody responses known to correlate with protection from severe disease across COVID-19 severities. Overall, our findings contribute to the evolving understanding of COVID-19 antibody dynamics.

**Funding:** Austin Public Health, NIAAA, Babson Diagnostics, Dell Medical School Startup.

## Introduction

Severe acute respiratory syndrome coronavirus 2 (SARS-CoV-2) infection, leading to Coronavirus disease 2019 (COVID-19), has accounted for more than 766 million infections and nearly 7 million deaths worldwide.^1^ Although SARS-CoV-2 vaccines have substantially lowered severe COVID-19 rates, only around 17% of the total US population has received the bivalent booster^2^ and the ongoing emergence of new variants, reinfections, vaccine hesitancy, and immune escape from monoclonal antibodies^3^ emphasize the continued need to study longitudinal immune responses to SARS-CoV-2 infection and vaccination.

The humoral immune response to SARS-CoV-2 infection or vaccination is multifaceted, leading to production of diverse antibodies to multiple epitopes that emerge with different kinetics and have distinct roles in immune protection.^4^ For example, antibodies to the nucleocapsid (N) protein are made early in the infection,^5^ do not have viral neutralizing capacity, and are generated only in response to the natural SARS-CoV-2 infection and not vaccination. Interestingly, anti-N antibodies may be generated due to cross-reactivity to other coronaviruses.^6^ In turn, antibodies to the receptor binding domain (RBD) of the spike (S) protein emerge slightly later post-infection, are responsible for viral neutralization, and are produced in response to both natural SARS-CoV-2 infection and vaccination, as currently approved vaccines encode S.^6^

While some studies suggest that more severe COVID-19 disease correlates with higher antibody titers to SARS-CoV-2,^7,8^ other studies found no differences in antibody levels between individuals with different illness severities.^9,10^ The difference in findings in the existing literature may relate to variation in study design, such as inclusion of small cohorts, focus on short-term follow-up of up to 6 months, lack of inclusion of asymptomatic individuals, or primary focus on either N, RBD, or S antibody dynamics. Also, few studies have directly compared SARS-CoV-2 antibody titers in the same cohort of individuals prior to and post-SARS-CoV-2 vaccination. Given that different SARS-CoV-2 antibodies serve different functions, it continues to be important to discern the time course that specific SARS-CoV-2 antibodies follow and whether these patterns vary in individuals with different initial peak disease severities. Understanding these temporal kinetics and longevity of specific SARS-CoV-2 antibodies will continue to inform strategies to minimize infectious surges and their severity, aid in vaccination and therapy approaches, and provide tailored clinical advice to patients on timing of vaccinations.

In this study, we quantified the dynamics and duration of anti-N, anti-RBD, and anti-S IgG antibody titers over the course of 12 months in a cohort of 98 participants with asymptomatic, mild, severe, and critical COVID-19 compared to titers in uninfected controls, utilizing four SARS-CoV-2 antibody assays. In addition, we report on antibody responses in relation to clinical parameters, such as age, sex, immunosuppressant use during acute COVID-19, and Post-Acute Sequelae of COVID-19 (PASC) status.

## Materials and Methods

### Institutional Review Board

This study was approved by the UT Austin Institutional Review Board (IRB ID: 2020-04-0117). Written informed consent was obtained from all participants in accordance with local and national regulations and all participant data were anonymized.

### Participant Recruitment

98 participants positive for a first acute SARS-CoV-2 infection and 17 healthy participant controls (controls) with no known COVID-19 exposure were recruited in Central Texas between July 2020 and September 2021 (Fig.1). Each participant’s disease severity was categorized as asymptomatic (no symptoms, non-hospitalized), mild (symptomatic, non-hospitalized), severe (hospitalized, non-intensive care unit), or critical (hospitalized, intensive care unit). For some analyses, participants were categorized by hospitalization status (asymptomatic and mildly ill (non-hospitalized); severely and critically ill (hospitalized)). COVID-19 status was confirmed by PCR or Atellica IM sCOVG assay (Ref. 11207388, Siemens Healthineers, Pennsylvania, USA). In addition to negative PCR and anti-SARS-CoV-2 antibody test status, healthy controls also reported no COVID-19 symptoms at the time of enrollment. Blood samples from COVID-19 participants were collected at four time intervals post-infection: 1, 3, 6, and 12 months following the initial positive SARS-CoV-2 test result. Participants were recruited within 2 weeks of each defined timepoint. A total of 8 specimens were collected outside of these defined time windows (Supp. Fig. 1) and were only included in the analyses when time was treated as a continuous variable. Samples from COVID-19 negative controls were collected once. EMR records were reviewed for steroid administration for downstream analysis.

**Figure 1.**
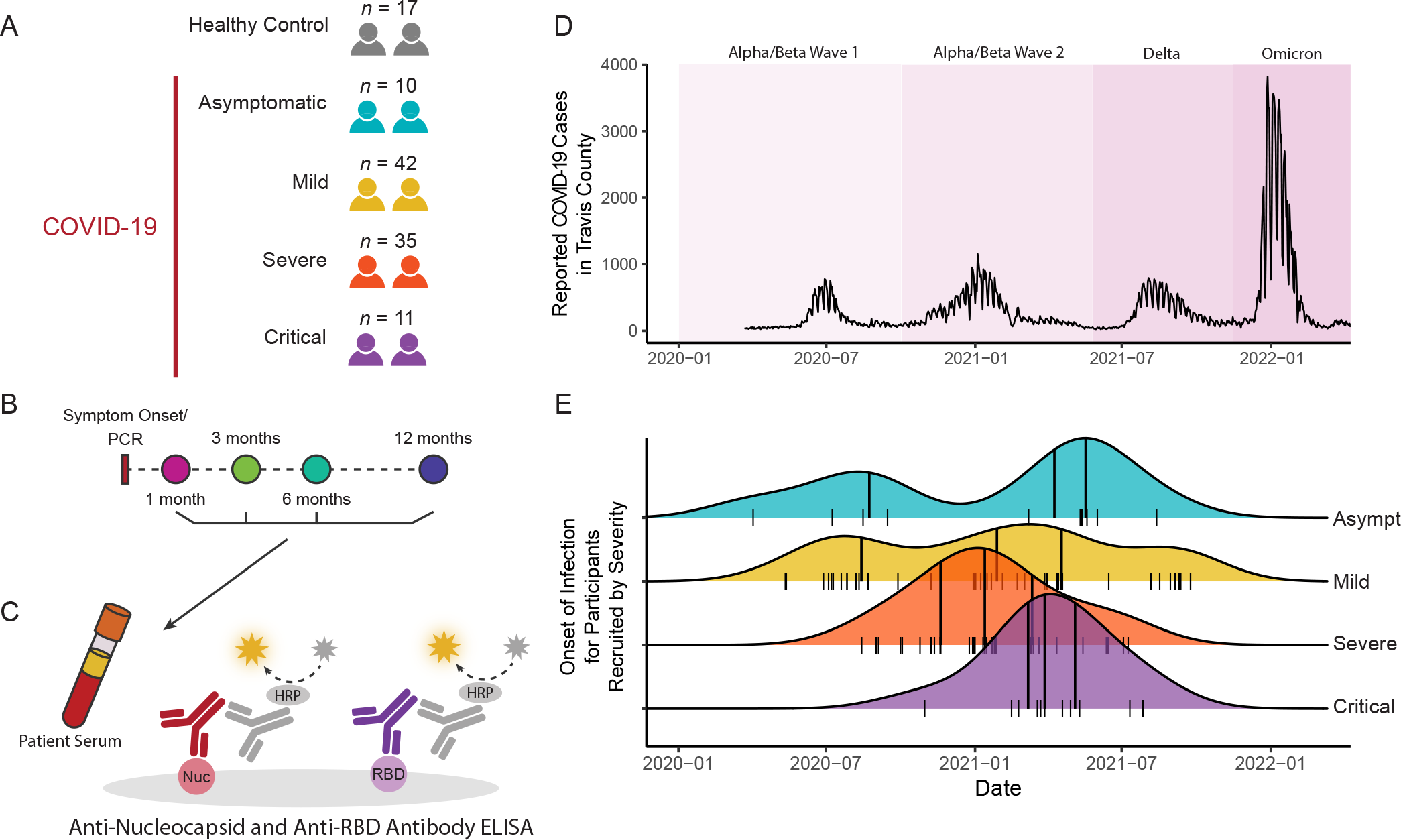
Participant Recruitment and Experimental Design. **A)** Sample size of acute COVID-19 participants in the cohort by disease severity, **B)** Participant serum was collected at 1, 3, 6, and 12 months post-positive PCR, **C)** ELISA schematic of detection of N and RBD antibodies in participant serum, **D)** Reported COVID-19 cases in the recruitment county (Travis County) over the time of recruitment, and **E)** Distribution of symptom onset or PCR+ for asymptomatic individuals in the cohort by acute COVID-19 severity. (Short tick marks = individual participants’ symptom onset or PCR+; Long tick marks = interquartile range within each disease severity group.)

### Sample Collection and Preparation

After the initial consent visit, participants were contacted via phone or email for sample collection at 1, 3, 6, and 12 months post-initial SARS-CoV-2 testing. Blood samples were collected in Ethylenediaminetetraacetic acid (EDTA) tubes (Greiner Bio-One Vacuette EDTA tubes cat. no. 456002 or BD Vacutainer EDTA tubes cat. No. 367899) at hospitals in the Austin area, at the UT Health Austin outpatient center, or in participant homes between July 2020 and September 2022. The samples, processed the same day of collection, were centrifuged at 2000 rpm at 4 degrees Celsius for 10 minutes and plasma was aliquoted and stored at -80 degrees Celsius for future immune assays (except for one participant, who had only serum banked for one timepoint). In addition to samples, participants were surveyed about vaccination status, reinfections, and symptoms at each visit.

### ELISAs

Participant plasma samples were tested using four SARS-CoV-2 assays. First, two commercial semi-quantitative enzyme-linked immunosorbent assays (ELISA) were used to detect IgG titers against SARS-CoV-2 N and RBD antigens (Cat. No. Nucleocapsid 448007, RBD 447707, BioLegend, California, USA). The ELISAs were run according to manufacturer’s protocol, except the S309 antibody,^11^ which binds the SARS-CoV-2 S protein, was substituted for the BioLegend-supplied anti-RBD standard antibody to ensure consistency between RBD and S assays. Second, Anti-RBD IgG titers were additionally measured using an indirect chemiluminescent immunoassay using the Atellica IM sCOVG assay (Ref. 11207388, Siemens Healthineers, Pennsylvania, USA) following manufacturer’s instructions. Finally, research ELISAs were developed to quantify IgG titers against whole SARS-CoV-2 S protein. Participant plasma samples for ELISAs were diluted at ratios 1:250, 1:750, 1:2250, 1:6750, and 1:20250 and run in duplicate. For ELISAs, two COVID-positive participant samples were included across multiple plates as internal batch controls, and results were measured using a FlexStation 3 Multi-Mode Microplate Reader (Molecular Devices, California, USA) at an optical density of 450 nm and 570 nm. The absorbance at 570 nm was then subtracted from the absorbance at 450 nm to remove background. Unless specified otherwise, anti-N and anti-RBD commercial ELISA results were used in the data analysis.

For the development of research ELISAs, HexaPro S protein^12^ was coated at 1 µg/mL on Costar® 96-well assay plates (High Binding polystyrene, Corning) overnight at 4 degrees Celsius. Plates were blocked with 5% milk in 0·1% Tween-20 in phosphate buffered saline (PBS-T) for 1 hour at room temperature. The S309 standard control antibody at 100 ng/mL was serially diluted three-fold seven times in PBS-T milk. Both participant samples and standards were run in duplicate. Antibody was allowed to bind for 45 minutes at room temperature, plates were washed three times with PBS-T, and goat anti-human Fc-HRP secondary (Southern Biotech) was added for 30 minutes at room temperature. After washing three times with PBS-T, Pierce TMB Substrate (Thermo Fisher) was added. HCl was then added to quench the colorimetric reaction and absorbance of each well at 450 nm was measured.

### Statistical Analysis

Antibody concentrations in ELISAs were quantified using standard curves. The standard curve data were modeled using the four-parameter logistic (4PL) curve-fitting method.^13^ The quality of the curve fit was assessed using the standard recovery procedure with recovery values between 70-120%.^14^ The 4PL equation was subsequently used to calculate unknown concentrations in the sample’s serial dilution set; the concentrations that best fit the curve were selected.

To model longitudinal antibody decay to natural infection exclusively, RBD data collected after a subject received a vaccination or was reinfected were excluded from analyses unless otherwise noted. To account for reinfections during the study, samples with anti-N and anti-RBD titers that were 1·5 times higher relative to their preceding titer were considered a reinfection, even if COVID-19 symptoms were not reported by the participant. Antibody titers were log10-transformed for modeling and statistical analysis (or for downstream analysis).

The association between increasing disease severity and antibody levels at different timepoints was measured using cumulative link models,^15^ while controlling for age and sex. For timepoints with a significant association, the Wilcoxon rank-sum test with Benjamin/Hochberg p-value correction was used for pairwise comparisons between disease severity groups were made using the Wilcoxon rank-sum test with Benjamini/Hochberg p-value corrections. Generalized additive mixed effect modeling with the R package mgcv (v 1.8.40) was used to evaluate the significance of severity in longitudinal antibody decay while controlling for age and sex.

Survival analysis was performed to estimate time to titers falling below the 95%-quantile-control-levels and time to titers falling below half of 1-month titers for all severities using the R package Interval (v 1.1.0.8).^16^ Rate parameters (lambdas) referred to as decay constants, were calculated based on the parametric exponential survival model. Our data were interval and right censored due to unknown exact time-to-event.

For all analyses except those in which pre- and post-vaccination titers were assessed, datapoints for anti-RBD and anti-S were excluded for samples collected after participant vaccination. Datapoints for antibody titers to all antigens were excluded post reinfection.

All graphing and analyses were conducted using R 4.1.0. and an alpha of 0·05 was used to determine significance across all statistical testing. Computational analyses were performed using the Biomedical Research Computing Facility at UT Austin, Center for Biomedical Research Support. RRID#: SCR\_021979.

## Data Availability

The anonymized data supporting this publication is available at ImmPort (https://www.immport.org) under study accession SDY2423.

## Acknowledgements

We are grateful to Hilary Selden for assistance with ordering supplies and Erica Brown for phlebotomy assistance. We appreciate Dell Medical School Neurology and Psychiatry department’s administrative support. This work was supported by Austin Public Health grant 4700 NI210000003 (S.S., E.M. L.I.R.E), NIAAA K08 T26-1616-11 (E.M.), NIH/NIAID R01AI104870 (L.I.R.E.), research funds from Babson Diagnostics (E.M.), and institutional Dell Medical School Startup funding (E.M.).

## Data Sharing

Data collected for the study, including de-identified individual participant data and a data dictionary defining each field in the set and related documents, such as the study protocol, will be made available to all with publication through ImmPort.

Additionally, all analysis codes will be made available to all with publication through Bitbucket.

## Results

### Participant Demographics

Ninety-eight participants with PCR-confirmed SARS-CoV-2 and 17 age- and sex-matched controls without history of COVID-19 infection or vaccination were enrolled into the study. Participants were stratified by self-reported COVID-19 severity, with 10 (10%) asymptomatic, 42 (43%) mildly ill (symptomatic, non-hospitalized), 35 (36%) severely ill (hospitalized, non-intensive care unit (ICU)), and 11 (11%) critically ill (hospitalized, ICU) participants. Of the COVID-19 positive cohort, 79 (81%) participants had biospecimens collected at 1 month, and 52 (51%) had biospecimens collected at all four timepoints (1, 3, 6, and 12 months), with an average of 3·6 samples per infected participant (Supp. Fig. 1). All samples (365) were assayed for anti-N and anti-RBD IgG titers, and 306 (84%) were assayed for anti-S IgG titers.

Participant ages spanned 18 to 87 years and the median age for the COVID-19 positive cohort was 42 years. 51 (52%) of the SARS-CoV-2 positive participants were female and 46 participants (47%) identified as Hispanic, with additional demographic and clinical information provided in Table 1.

**Table 1.**
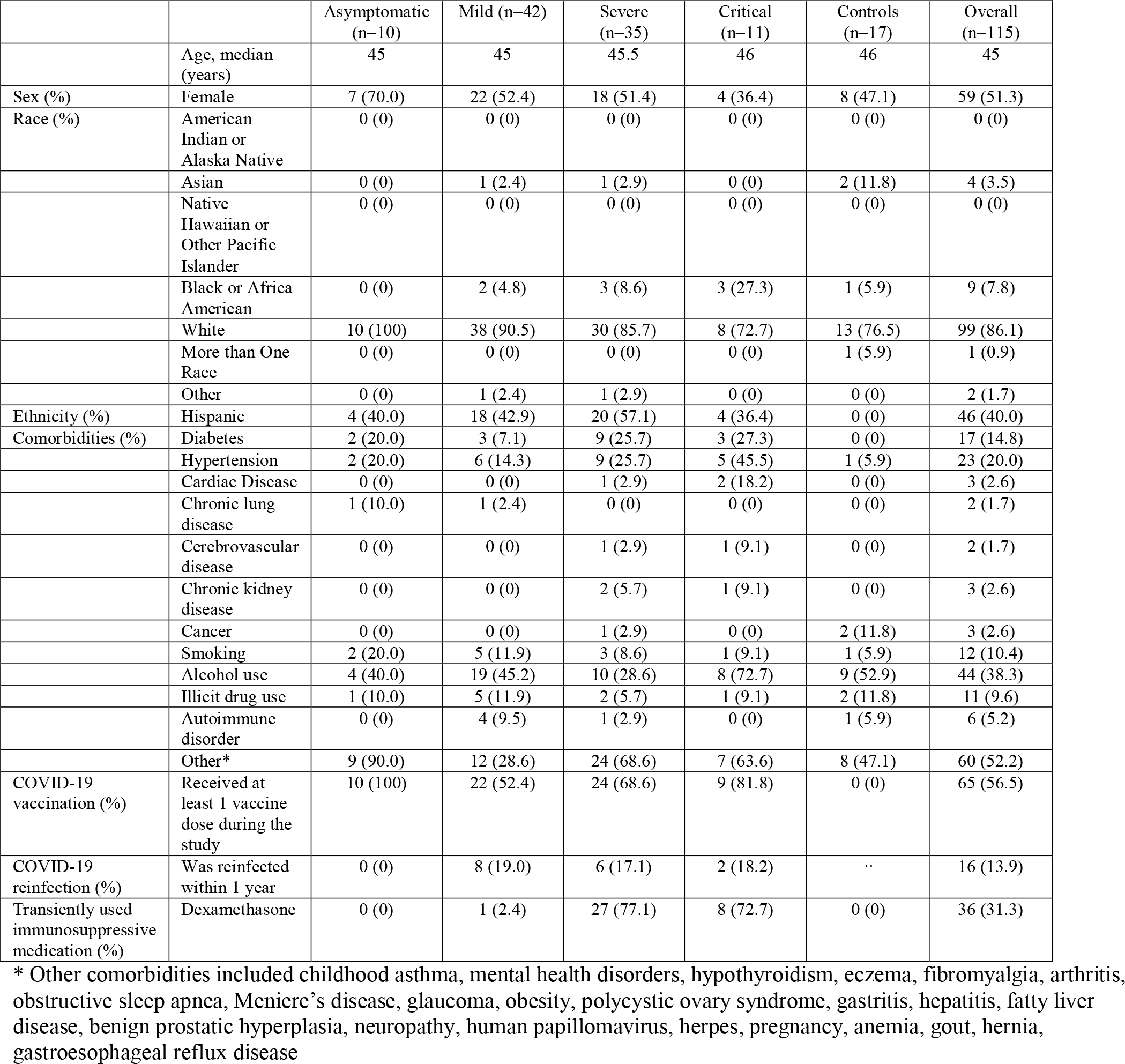
Patient Demographics.

Enrollment in the study was restricted to unvaccinated participants. Between their positive PCR test and the end of the study, 65 (57%) participants received at least one COVID-19 vaccination. 16 (14%) participants experienced reinfections during the study, determined by either participant reporting a reinfection or a 1·5-fold increase in anti-N and anti-RBD titers relative to the prior sample collection timepoint.

### Concordant Antibody Measurements Across Laboratory-Developed and Commercial Assays

We analyzed anti-SARS-CoV-2 titers across four different assays and measured associations between antibody responses to RBD versus whole S protein at 1, 3, 6, and 12 months post-acute SARS-CoV-2 infection for participants with different disease severities. Anti-RBD titers, assayed with a commercial ELISA kit, were comparable to anti-RBD titers assayed with the Siemens large-scale indirect chemiluminescent immunoassay (Supp. Fig. 2A, Spearman correlation R = 0·54, p < 2·2e-16). Anti-RBD titers also strongly correlated with anti-S titers by ELISA, validating the anti-S laboratory-developed ELISA (Supp. Fig. 2B Spearman correlation R = 0·83, p < 2·2e-16).

### Anti-SARS-CoV-2 Antibody Titers Correlate with Acute COVID-19 Severity Both Initially and in Convalescence

We next sought to determine whether there was a difference in convalescent SARS-CoV-2 antibody titers based on acute COVID-19 severity. We found that anti-N, anti-RBD, and anti-S IgG titers were significantly associated with acute disease severity at 1 month post-positive SARS-CoV-2 PCR (overall anti-N p.adj = 1·3E-4, anti-RBD p.adj = 8·5E-3, anti-S p.adj = 8·5E-3, controlling for age and sex) (Fig. 2A-C), with the critically ill participants exhibiting the highest convalescent antibody titers. Notably, the association between acute disease severity and convalescent antibody titers was maintained over 12 months for anti-N IgG (3 months p.adj = 0·0012, 6 months p.adj = 0·0021, 12 months p.adj = 0·01), and for 3 months for anti-RBD IgG (3 months p.adj = 0·0094). While overall acute disease severity was significantly associated with anti-N titers up to 12 months, pairwise comparisons between severity groups at 12 months did not demonstrate statistical significance post-correction for multiple testing. Notably, we had lower power to resolve persistent association between severity and titers for anti-RBD and - S compared to anti-N because post-vaccination data were removed for these 2 assays.

**Figure 2.**
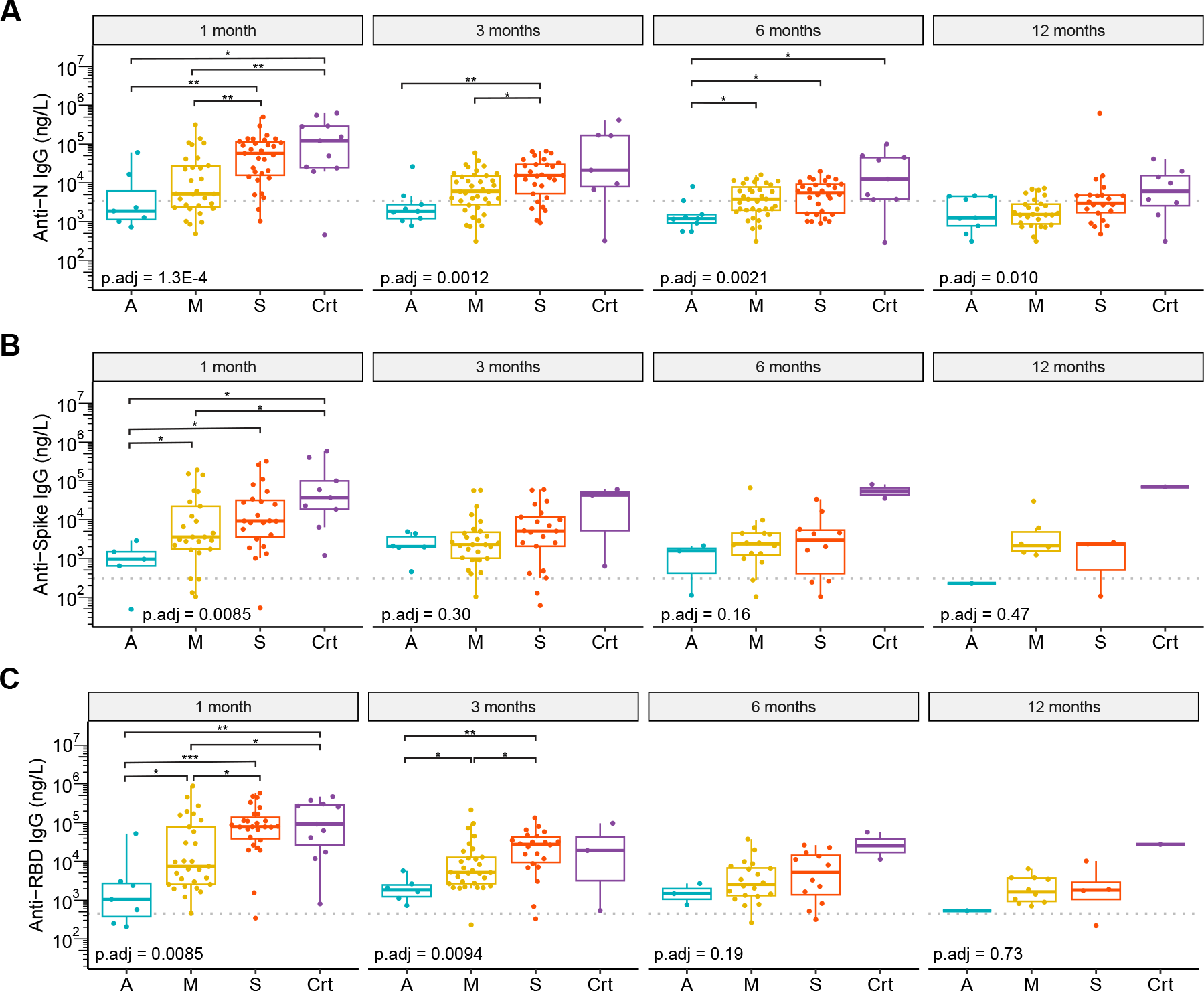
SARS-CoV-2 antibody titers correlate with COVID-19 disease severity. Box and whisker plots of **A)** Anti-N, **B)** Anti-S, and **C)** Anti-RBD IgG titers for asymptomatic (A), mild (M), severe (S), and critical (Crt) groups at 1, 3, 6, and 12 months post-positive SARS-CoV-2 PCR. (Global p value obtained with cumulative link model testing between disease severity and antibody titers, controlling for age and sex. Pairwise comparisons performed using Wilcox test, with FDR correction. * p.adj ≤ 0·05, ** p.adj ≤ 0·01, *** p.adj ≤ 0·001. Dotted lines indicate 95% quantile of healthy controls.)

### Vaccination Increases Anti-RBD Titers in Hospitalized and Non-Hospitalized Participants, but Decreases the Ratio of Anti-RBD:Anti-S

As antibodies against SARS-CoV-2 RBD are associated with viral neutralization, we sought to test the extent to which vaccination increased participants’ anti-RBD antibody titers. We evaluated anti-N and anti-RBD titers at 3, 6, and 12 months post-positive SARS-CoV-2 PCR, relative to participants’ titers at 1-month post-infection. Participant samples were stratified by hospitalization status, and we compared mean antibody titers in samples categorized by vaccination status (i.e., before and after vaccination). In pre-vaccination samples from both hospitalized and non-hospitalized participants, a decline in anti-N and anti-RBD levels is evident over the 12 month time-course (Fig. 3A-B). As expected, anti-N titers were not impacted by vaccination (Fig. 3A). Anti-RBD titers increased significantly following vaccination, regardless of participants’ hospitalization status (non-hospitalized: p = 1·5E-6 (3 months), p = 5·4E-6 (6 months), p = 3E-4 (12 months) and for hospitalized: p = 3E-3 (3 months), p = 3·9E-7 (6 months), p = 3E-3 (12 months)) (Fig. 3B). Notably, anti-RBD titers were elevated to a greater extent after vaccination in non-hospitalized compared to hospitalized participants at 3- (p.adj = 4E-2) and 12-month (p.adj = 4·7E-2) timepoints.

**Figure 3.**
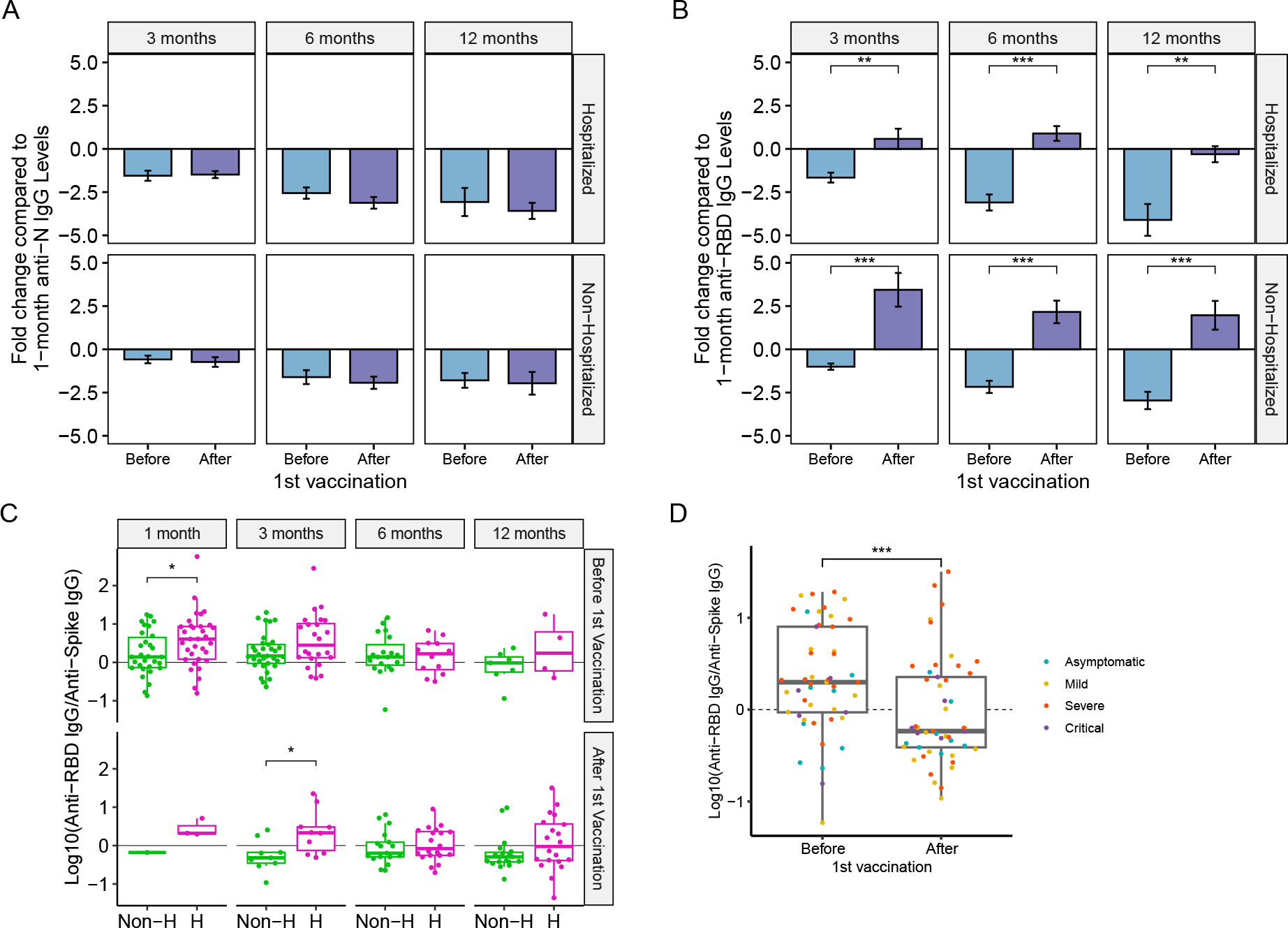
SARS-CoV-2 vaccination increases anti-RBD titers to a greater extent in participants who experienced milder disease but reduces anti-RBD:anti-S titers. Log2 fold change of **A)** Anti-N and **B)** Anti-RBD IgG titers relative to participants’ 1-month titers. **C)** Anti-RBD to anti-S ratio at 1, 3, 6 and 12 months post-positive SARS-CoV2 PCR, separating samples based on whether they occurred before and after 1st vaccination, and **D)** Anti-RBD to anti-S IgG ratio before and after 1st vaccination for all samples. Pairwise comparisons performed using Wilcox test, with FDR correction. * p.adj ≤ 0·05, ** p.adj ≤ 0·01, *** p.adj ≤ 0·001. (Dashed lines indicate 1-month titer levels)

Considering the essential role of anti-RBD in viral neutralization, we next tested if the relative titers of antibodies against RBD versus total S protein differed according to hospitalization or vaccine status. Prior to vaccination, hospitalized participants had a significantly higher anti-RBD to anti-S IgG ratio at 1-month post-PCR test compared to non-hospitalized participants (p.adj = 0·048). Similarly, post-vaccination, hospitalized participants had an elevated ratio of anti-RBD to anti-S IgG at 3 months post-PCR test relative to non-hospitalized participants (p.adj = 0·017) (Fig. 3C). In addition, paired comparisons of mean titers pre- and post-first vaccination across all disease severities demonstrated that the anti-RBD to anti-S IgG ratio significantly decreased in convalescent participants post-vaccination (p = 1·1e-4) (Fig. 3D).

### Anti-N SARS-CoV-2 Antibody Titers Decay Faster, But Remain Higher, in Hospitalized Compared to Non-Hospitalized Participants Over 12 Months Post-SARS-CoV-2 Infection

We next analyzed the decay kinetics of anti-N IgG titers (for all samples) and anti-RBD IgG titers (for pre-vaccination samples) over the course of 12 months post-confirmed infection. Anti-N and anti-RBD titers declined over 12 months for participants of all disease severities (Figure 4A-B). However, the rate of decay was generally greater in hospitalized relative to non-hospitalized participants. Specifically, as a measure of antibody longevity, we examined the percent of participants at 3, 6, and 12 months who sustained SARS-CoV-2 antibody titers against N and RBD above 50% of their individual 1-month levels (Figure 4C-D). The proportion of participants with anti-N titers remaining above 50% of their 1-month levels declined with decay constant of 0.604 month^-1^ in hospitalized versus 0.237 month^-1^ non-hospitalized participants, and anti-RBD titers declined with decay constants of 0.628 month-1 in hospitalized versus 0.345 month-1 non-hospitalized participants. Of note, despite the faster antibody decay rate in hospitalized participants, given their substantially higher anti-N and anti-RBD titers at 1 month, antibody titers generally remained higher over the study period in hospitalized relative to non-hospitalized participants (Figure 4A-B; overall anti-N p.adj = 1·3E-4, anti-RBD p.adj = 8·5E-3, controlling for age and sex).

**Figure 4.**
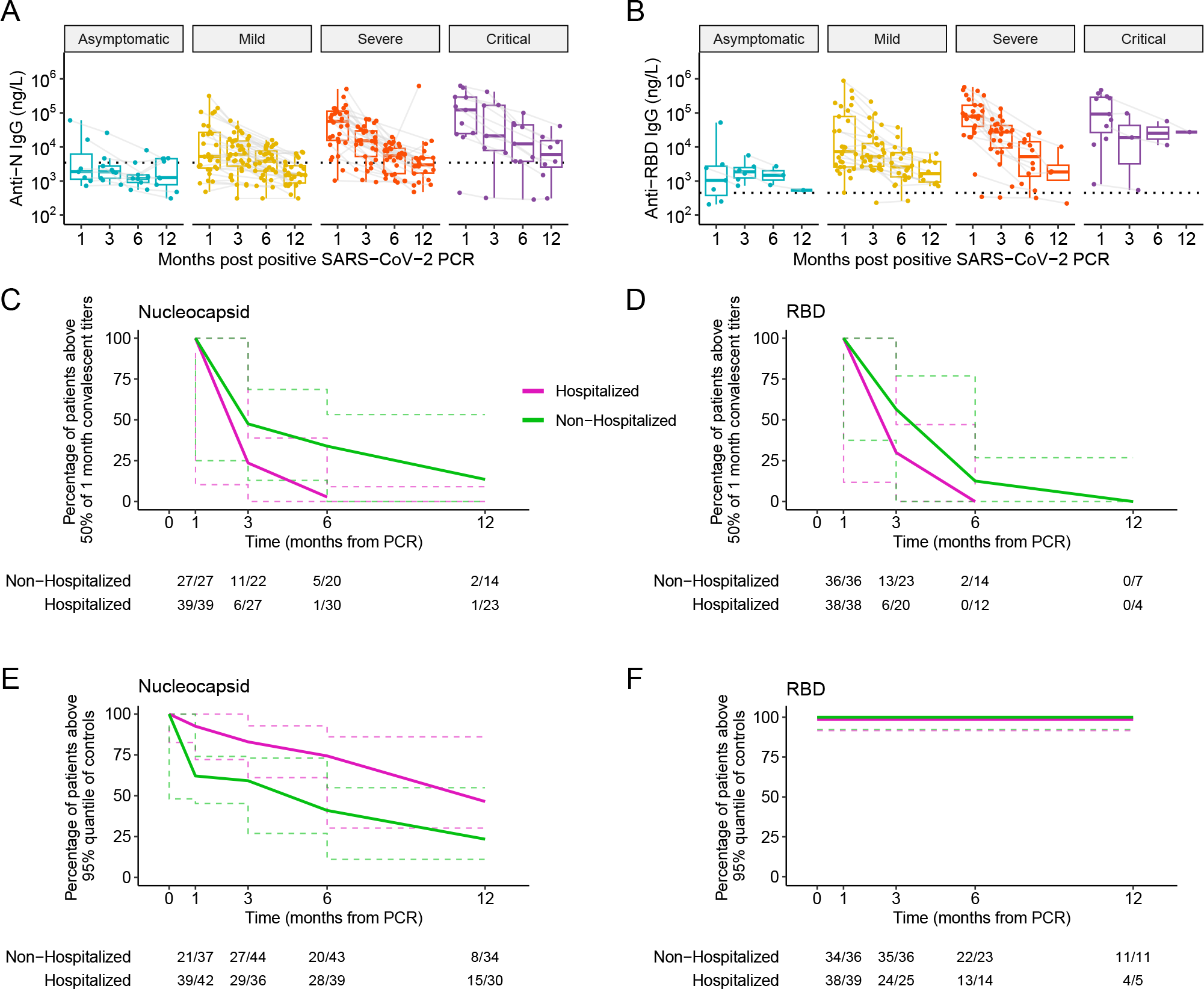
Anti-N titers decline below, while anti-RBD titers are sustained above control levels over 12 months, and anti-N IgG titers decay faster in hospitalized individuals. Longitudinal decay of **A)** anti-N and **B)** anti-RBD antibody titers in participants stratified by initial disease severity. Participant timepoints are connected by light gray lines. Control 95% confidence interval represented by dashed line. Percentage of participants whose **C**) anti-N titers and **D**) anti-RBD titers remained above 50% of their respective 1-month post-infection titers over 12 months. Percentage of participants whose **E)** anti-N and **F)** anti-RBD antibody titers remained above the 95^th^ quantile of controls over 12 months in all participants. Dashed lines in **C-F** represent 95% confidence intervals.

For anti-N, there was a significant difference in the frequencies of non-hospitalized versus hospitalized participants retaining titers above 50% of initial titers (asymptotic logrank two-sample test p value = 0·01). By 3 months, 50% of non-hospitalized and 22·2% hospitalized participants retained at least 50% of their 1-month anti-N titers (Fig. 4C). By 6 months, only 25% of non-hospitalized and 3·3% of hospitalized participants retained anti-N titers above 50% of 1-month levels, consistent with a faster antibody decay rate for hospitalized participants. For anti-RBD, by 3 months, 56·5% of non-hospitalized and 30% hospitalized participants had titers above 50% of their 1-month titers (Fig. 4D). This percentage decreased to 14·3% of non-hospitalized and 0% of hospitalized participants by 6 months and to 0% by 12 months for both non-hospitalized and hospitalized participants, indicating that none of the participants retained 50% of their initial RBD by 12 months of infection. However, there was no significant difference in the rate of decline in anti-RBD titers between non-hospitalized and hospitalized participants.

#### Longevity Of Anti-SARS-Cov-2 Antibody Titers in Participants Relative to Uninfected Control Participant Titers

To evaluate when anti-SARS-CoV-2 antibody titers decline to levels of uninfected control participants, we compared anti-N and anti-RBD IgG titers in infected participants over 12 months post-confirmed infection relative to the 95% quantile of titers in uninfected control participants (Fig. 4E-F). We observed that the frequency of participants with anti-N titers over control levels declined faster for non-hospitalized compared to hospitalized participants (asymptotic logrank two-sample test p value = 0·002). At 1 and 3 months post-infection, over 50% of non-hospitalized and hospitalized participants had antibody titers exceeding the 95% quantile of control levels (Fig. 4E-F). By 6 months, 46·5% of non-hospitalized and 71·8% of hospitalized participants maintained anti-N antibody levels above control titers. Finally, by 12 months, less than 25% of non-hospitalized and 50% of hospitalized participants retained anti-N antibody levels above control titers, demonstrating a faster decline in the frequency of non-hospitalized participants whose anti-N titers remain above control levels. Interestingly, although anti-RBD titers decayed with time (Fig 4B), they remained above the 95% quantile of pre-COVID-19 control levels over 12 months, regardless of acute disease severity (Fig. 4F).

### Age Correlates with SARS-CoV-2 Antibody Titers at Early Post-Infection Timepoints

There was a moderate, but significant positive correlation between age and antibody titers at 1, 3, and 6 months for anti-N (Spearman correlation R = 0·32, p = 0·0049 (1 month), R = 0·26 with p = 0·017 (3 months), R = 0·23 with p = 0·037), and at 1 month for anti-RBD antibodies (Fig. 5A-B, Spearman correlation R = 0·34 with p = 0·0041). These correlations remained significant despite controlling for severity. In contrast, there was no correlation between sex and antibody titers at any of the tested timepoints (Fig. 5C-D).

**Figure 5.**
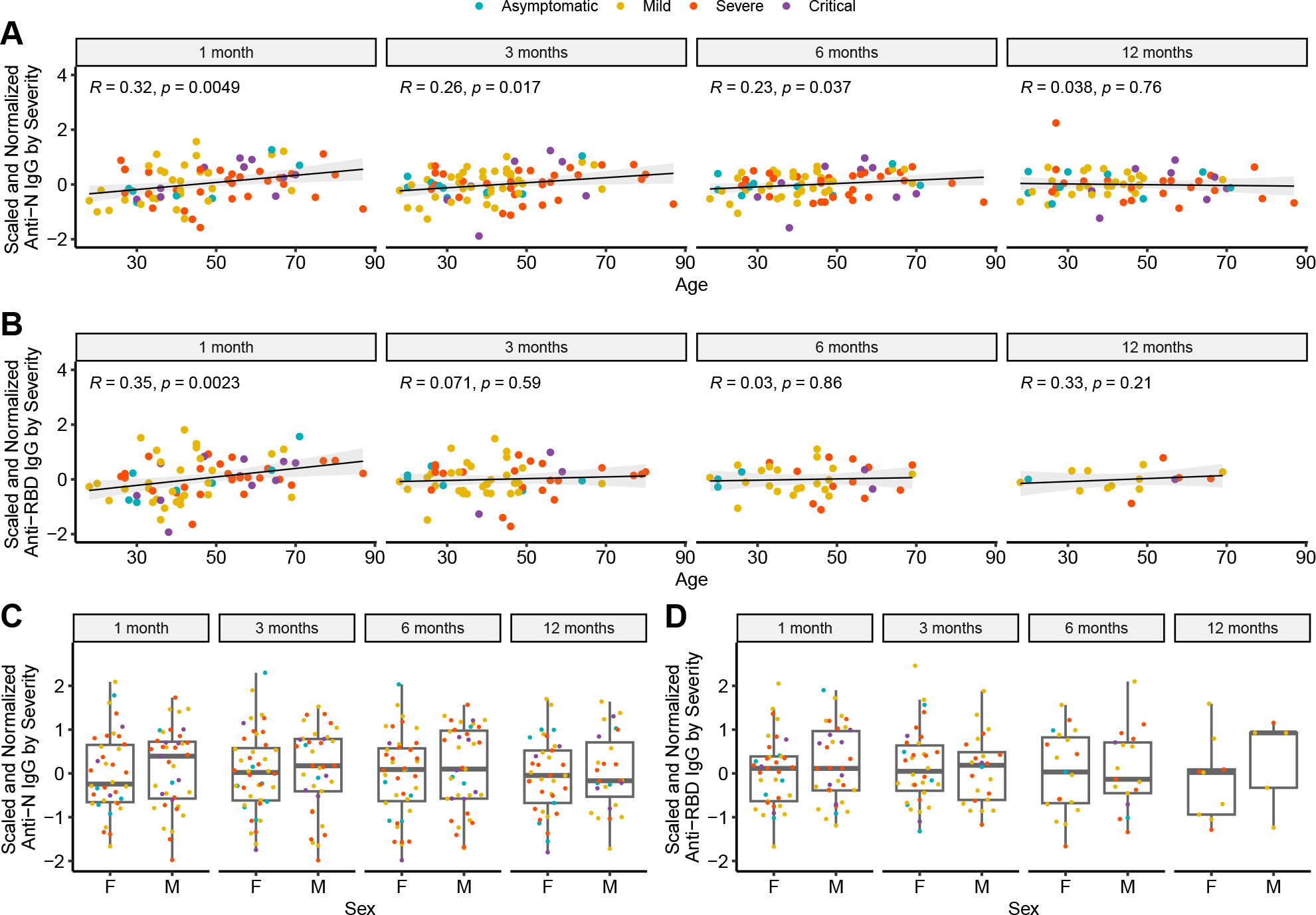
Age correlates with SARS-CoV-2 antibodies in convalescent participants. Correlation between age and **A)** anti-N and **B)** anti-RBD IgG, and between sex and **C)** anti-N and **D)** anti-RBD IgG while controlling for disease severity (F = female, M = male).

### PASC Status is Not Associated with Anti-N and Anti-RBD Convalescent Antibody Titers

In our study, participants who had at least one persistent symptom per body system for longer than 1 month after a positive COVID-19 test at 2 or more visits were classified as having Post-Acute Sequelae of COVID-19 (PASC). Using this criterion, 16·3% of the cohort developed PASC, which included long-term symptoms such as brain fog, fatigue, insomnia, headaches, myalgias, cough, and dyspnea (Table 2). Stratifying participants by disease severity groups, 0/10 (0%) of asymptomatic, 6/42 (14·3%) of mildly ill, 8/35 (22·9%) of severely ill, and 2/11 (18·2%) of critically ill participants reported PASC symptoms. There was no significant correlation between PASC status and anti-N or anti-RBD antibody titers at initial or subsequent timepoints over the course of 12 months.

**Table 2.**
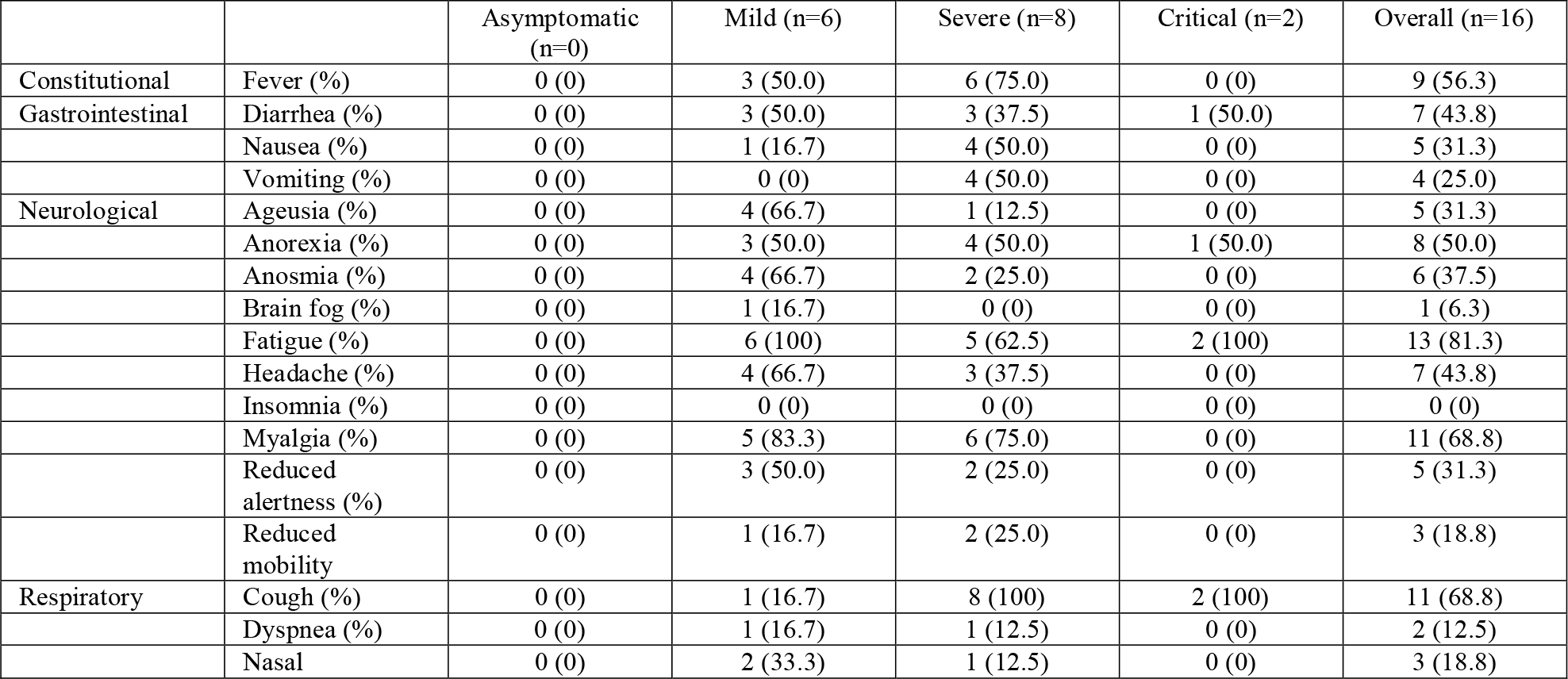
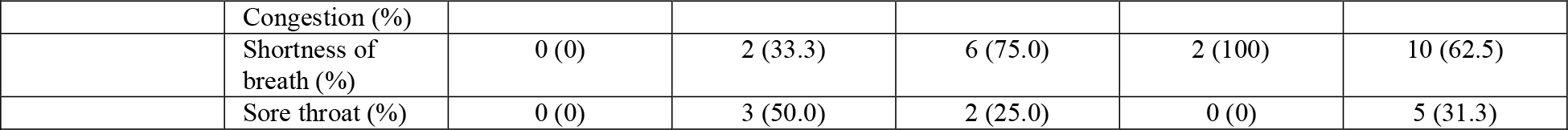
PASC Symptoms by Acute COVID-19 Severity.

### No Association Between Acute Stage Immunosuppression Treatment and Convalescent Anti-N and Anti-RBD Antibody Titers

We also evaluated whether treatment with steroids in acute infection affected anti-N and anti-RBD antibody titers over the course of a year in hospitalized participants. Although there was a trend for higher anti-N and anti-RBD antibodies across 1, 3, and 6 months post-infection in participants treated with steroids, statistical significance was not reached (Fig. 6).

**Figure 6.**
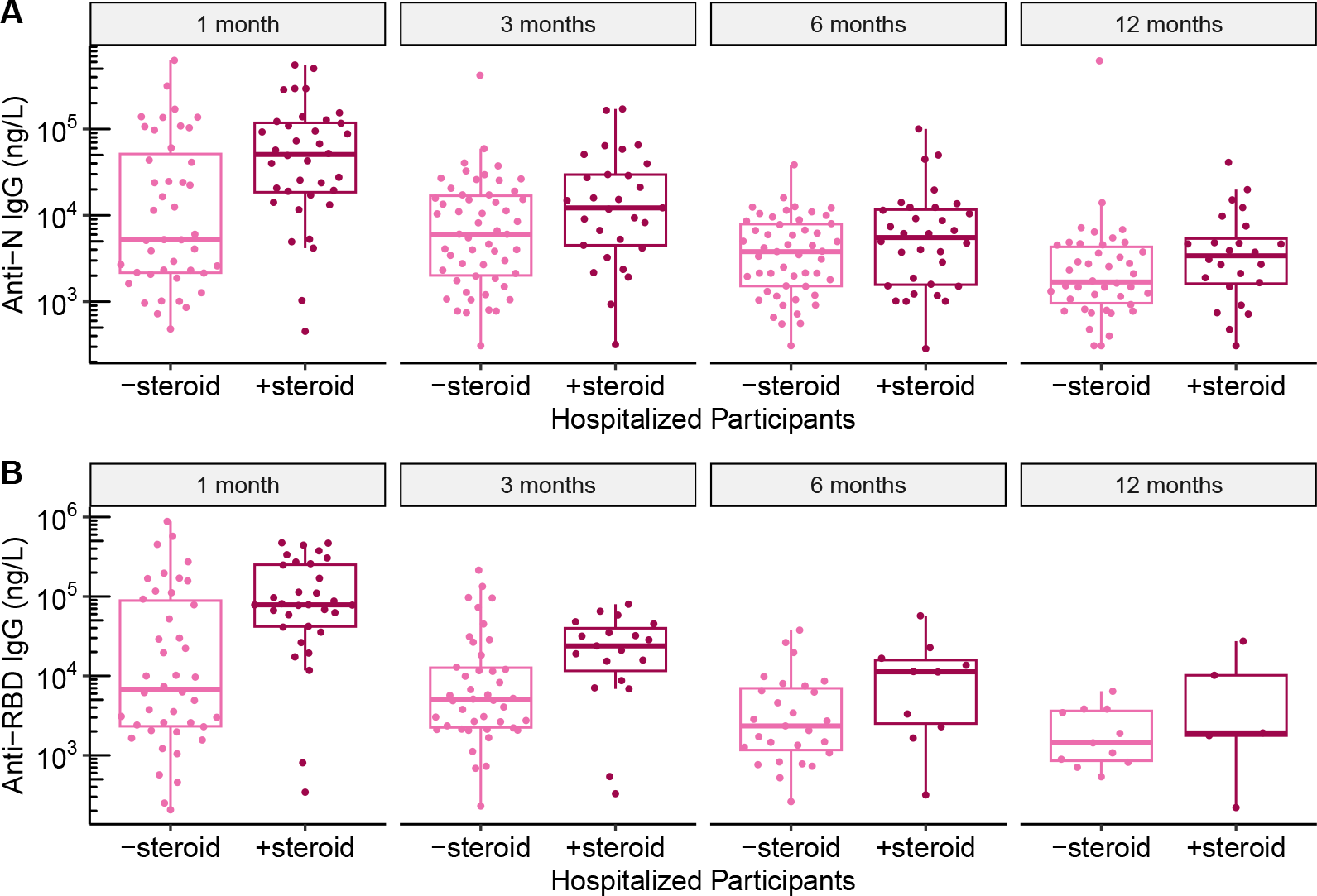
Steroid treatment did not correlate with SARS-CoV-2 antibody titers in hospitalized participants. **A)** anti-N and **B)** anti-RBD IgG titers in hospitalized participants across 1, 3, 6, and 12 months post-SARS-CoV-2 infection, stratified by treatment with steroids (Wilcox test, p>0·05 for all pairwise comparisons of titers in participants treated with or without steroids).

## Discussion

Despite the advent of vaccines, SARS-CoV-2 continues to be a global threat to public health, with several hundred individuals still being hospitalized for and dying of COVID-19 each week in the United States^17^ as well as continued resurgence of new variants leading to reinfections. Understanding the longevity of SARS-CoV-2 antibody responses is crucial for assessing the likelihood of reinfections and the efficacy as well as timing of vaccinations. In this study, we used four different assays to investigate the antibody responses to SARS-CoV-2 N, RBD, and S proteins in 98 participants compared to 17 uninfected controls. Our analysis offers insights into SARS-CoV-2 antibody dynamics in individuals with different acute disease severities over 12 months post-COVID-19 infection with potential to inform vaccination strategies in the general population.

As specific SARS-CoV-2 antibodies serve different roles, their individual characterization is necessary to fully evaluate the SARS-CoV-2 immune response. For example, anti-N antibodies remain the best marker of natural infection as N is not currently included in vaccines used in the United States.^18^ In turn, anti-RBD antibodies, a subset of anti-S antibodies, are important for viral neutralization and inhibition of SARS-CoV-2 infection by blocking either the ACE2-RBD binding interactions or S-mediated membrane fusion.^19,20^ SARS-CoV-2 neutralizing antibody titers correlate with total anti-S and anti-RBD serum titers, and increasing titers of both neutralizing and total binding antibodies correlate with protection from disease.^21^ In fact, a threshold of anti-RBD titers has been identified that correlates with multiple protective activities of humoral immunity.^22^ In our study, we found that anti-N, anti-S, and anti-RBD antibodies at 1 month post-infection were higher in severely and critically ill participants compared to mildly ill and asymptomatic participants. This significant trend of higher antibody titers in hospitalized versus non-hospitalized participants persisted over the 12 months of this study.

The observed association of disease severity with higher SARS-CoV-2 antibody titers is in line with other studies^7,8^ and may be attributed to severely ill individuals having a higher viral load, resulting in elevated B cell activation.^23^ There are several possible explanations for the failure of higher anti-RBD titers, which are generally a correlate for protection from COVID-19, to protect hospitalized patients from experiencing more severe disease. First, anti-RBD antibodies may have arisen too late to provide adequate protection against rising viral titers in patients with more severe disease,^24^ although these higher viral titers ultimately stimulate a strong antibody response. Other studies suggest that the functional quality of antibodies may better predict immune protection than the absolute quantity of antibodies,^25–27^ as the serum repertoire may be dominated by a few clonotypes, which can vary in their ability to mediate neutralization or other antibody-mediated effector functions.^28^ Consequently, severe patients may generate higher titer, but less protective antibody responses based on the SARS-CoV-2 epitopes recognized, the affinity of the antibodies for these epitopes, or their Fc effector functions, such that severe patients may require higher antibody titers to be protected from infection.

As COVID-19 transitions to an endemic disease, determining the minimal level of SARS-CoV-2 antibodies necessary to prevent reinfection and severe disease, as well as identifying the optimal vaccination schedule to maintain these levels, are of primary public health concern. Many studies have demonstrated that antibody-mediated immunity protects against SARS-CoV-2 reinfections up to 7 months post-infection.^29,30^ Interestingly, while both hospitalized and non-hospitalized individuals in our study developed higher anti-RBD titers post-vaccination, we observed a greater increase in RBD titers in non-hospitalized participants compared to hospitalized participants post-vaccination. This difference may reflect that non-hospitalized participants had lower viral loads that did not elicit as strong of a B cell response. The greater increase in antibody titers observed post-vaccination in non-hospitalized versus hospitalized participants suggests that asymptomatic/mild individuals may benefit even more from vaccination compared to severely ill individuals in the first few months after acute illness. Given that only 17% of the general population is currently vaccinated with the updated booster,^2^ our data raise the suggestion that mildly ill individuals may benefit the most from vaccinations to increase antibody titers and may benefit from vaccination earlier after infection due to lower anti-RBD titers starting as early as 1-month post positive PCR test. Interestingly, we found that while vaccination boosted anti-RBD titers in participants of all severities, the anti-RBD:anti-S ratio declined, suggesting that vaccines could be further improved to elicit a higher frequency of neutralizing antibodies.

Khoury and colleagues recently modeled neutralizing antibody titers relative to reported protection from multiple vaccine studies to suggest that just 3% of mean convalescent antibody levels were sufficient to protect against severe infection.^31^ This finding suggests that low levels of antibodies are sufficient to protect against severe disease and mitigate reinfections, likely because antibodies provide immune protection through mechanisms beyond neutralization, such as opsonization and antibody-dependent cellular cytotoxicity. Our study extends prior research, demonstrating that anti-RBD levels remain above control levels up to 12 months post-infection for participants across acute disease severities. Although less than 12% of participants in our cohort retained 50% of their initial RBD antibody titers beyond 6 months post-infection, and none retained 50% titers at 12 months post-infection, we find that anti-RBD levels remain above control levels up to 12 months post-infection for participants across acute disease severities. These findings suggest that most convalescent individuals retain sufficient antibody titers for protection against severe infections for up to 12 months after infection. Interestingly, antibody titers in participants with more severe disease decayed faster than in participants with mild disease. While the basis for this difference is not known, severe patients may have fewer circulating plasmablasts entering the long-lived plasmablast compartment, resulting in more rapid decay of serum titers. Alternatively, antibodies in severe patients may be cleared from circulation more rapidly due to reduced FcRn-mediated recirculation or increased Fc-mediated clearance from circulation. Despite their faster antibody decay rate, participants who experienced severe acute COVID-19 disease had such high titers at 1 month that they retained higher levels of anti-N and anti-RBD antibody titers over the course of 12 months compared to participants who experienced mild disease.

Notably, even with waning serum antibody levels over time, other parts of the immune system could help protect individuals from reinfections. For example, upon re-exposure to SARS-CoV-2, the synthesis of IgA antibodies in the mucous membranes of the respiratory tract may aid in preventing or impeding SARS-CoV-2 infection.^32^ Furthermore, memory T cells and NK cells can also provide effective anti-viral memory immune responses mitigating reinfection.^33^ Previously, we reported that individuals with multiple sclerosis who were depleted of B cells, do not mount strong antibody responses, but also do not generally develop severe disease,^34^ indicating that other arms of the immune system are protective and may be able to prevent reinfection, even when antibody responses are very minimal. Nevertheless, our data suggest that timing of vaccination schedules should be optimized between 6-12 months post-infection and should account for individuals’ initial disease severity, as more severe acute disease correlates with longer persistence of elevated antibody titers.

Beyond the humoral and cellular immune system, immune responses to infections are also influenced by multiple other factors, such as age, sex, comorbid conditions, and administered medications. Age influences B cell physiology, leading to decreased B cell production in the bone marrow and fewer memory B cells in the elderly.^35^ Older age and male sex have been associated with higher susceptibility to severe acute COVID-19 disease,^36^ although less is known about the relationship between these demographic factors and convalescent anti-SARS-CoV-2 antibody titers. While some studies have found higher convalescent SARS-CoV-2 antibodies in association with older age and male sex,^37^ others noted an association with younger age and female sex.^38^ Our results indicate a moderate positive age correlation with 1-month anti-RBD and 1- and 3-month anti-N titers, and we did not observe an association between sex and convalescent titers. It is possible that the age-antibody correlation may partially be driven by higher disease severity in older participants. Although our analyses adjusted for binned disease severity (i.e. asymptomatic, mild, severe, critical), it is possible that a positive correlation arises because older participants likely comprise more severe cases in each severity category, thus possibly rendering the adjustment of binned severity insufficient to fully decouple its effect from age. Nonetheless, it is interesting to note that despite the known decline in B-cell activity that occurs with age, older individuals in our study mounted robust antibody responses although the elevated titers post-infection were not sufficient to protect from severe disease. Further research is needed to clarify how different demographic factors may impact an individual’s humoral immune response post-COVID-19.

PASC has emerged as a multi-system disease in a subset of patients with COVID-19, with symptoms lasting for weeks to months following recovery from acute COVID-19 infection and leading to significant functional disability in affected individuals.^39^ In our cohort, 16·3% of the individuals experienced PASC, a comparable rate to other studies citing 10% to 60% of individuals experiencing PASC following acute COVID-19.^39,40^ We found no correlation between PASC status and anti-N or anti-RBD antibody titers at any timepoint, suggesting that the magnitude of convalescent antibodies may not be predictive of the likelihood of developing PASC. These results also support findings from other studies demonstrating that initial antibody titers or vaccine boosters did not affect PASC outcomes.^41^

Another factor that could potentially influence convalescent antibody levels is immunosuppressant use during acute stage of COVID-19, as steroids are known to suppress plasma cell differentiation.^42^ Steroids became the cornerstone of treatment early during the pandemic.^43^ Although steroids effectively reduce the excessive inflammatory response during the initial stages of COVID-19, their impact on the development of long-term SARS-CoV-2 immunity remains uncertain. In our study, no significant associations were found between RBD and N titers post-infection and steroid use in acute infection for hospitalized participants, though future studies with larger cohorts of individuals receiving steroids are needed to validate our finding that steroid administration does not influence convalescent COVID-19 titers.

The strengths of our study include a large cohort of both vaccinated and unvaccinated COVID-19 positive individuals studied longitudinally over 12 months with inclusion of COVID-19 negative controls; a high retention rate across multiple sample collections over the course of one year; inclusion of participants who were asymptomatic or had mild, severe, or critical disease; the ability to measure longitudinal antibody titers to natural infection versus vaccination; and use of four different assays to measure antibody titers for key SARS-CoV-2 antigens. Our study also had limitations. First, the cohort was primarily comprised of adult individuals in one geographic location, and we did not evaluate pediatric individuals. Second, our study did not assess specific SARS-CoV-2 variants, limiting our ability to generalize evaluation of antibody titer dynamics in response to evolving SARS-CoV-2 variants. Finally, we did not directly examine neutralizing titers or cellular mediated immunity, which are both critical aspects of SARS-CoV-2 immune responses.

In conclusion, our study characterizes the dynamics of anti-N, RBD, and S antibodies in individuals following SARS-CoV-2 infection, while considering their disease severity and vaccination history. We found a significant association between acute COVID-19 severity and the levels of SARS-CoV-2 antibodies in the convalescent period over the course of 12 months. As expected, convalescent anti-N antibodies decreased irrespective of vaccination, while anti-RBD antibodies increased post-vaccination in both hospitalized and non-hospitalized participants, with a higher increase in those who were non-hospitalized. Higher antibody decay rates pre-vaccination were observed in hospitalized relative to non-hospitalized individuals, despite the fact that hospitalized individuals maintained higher anti-N and anti-RBD titers over the course of 12 months. Only a small fraction of participants of any disease severity retained 50% of their initial antibodies at 6 months. Interestingly, while at 12 months, less than half of all participants maintained anti-N antibody levels above controls, anti-RBD levels remained consistently higher than control levels in all participants regardless of initial disease severity, potentially providing protection against severe reinfections.^31^ While there was a moderate age correlation with early-stage antibody levels, neither sex, PASC status, or acute immunosuppression treatment affected convalescent antibody levels. Overall, our findings contribute to the evolving understanding of SARS-CoV-2 antibody dynamics.

