## Supplemental Figure 1 for "SARS-CoV-2 Humoral Immune Responses in Convalescent Individuals Over 12 Months Reveal Severity-Dependent Antibody Dynamics"

**Supp. Fig. 1**

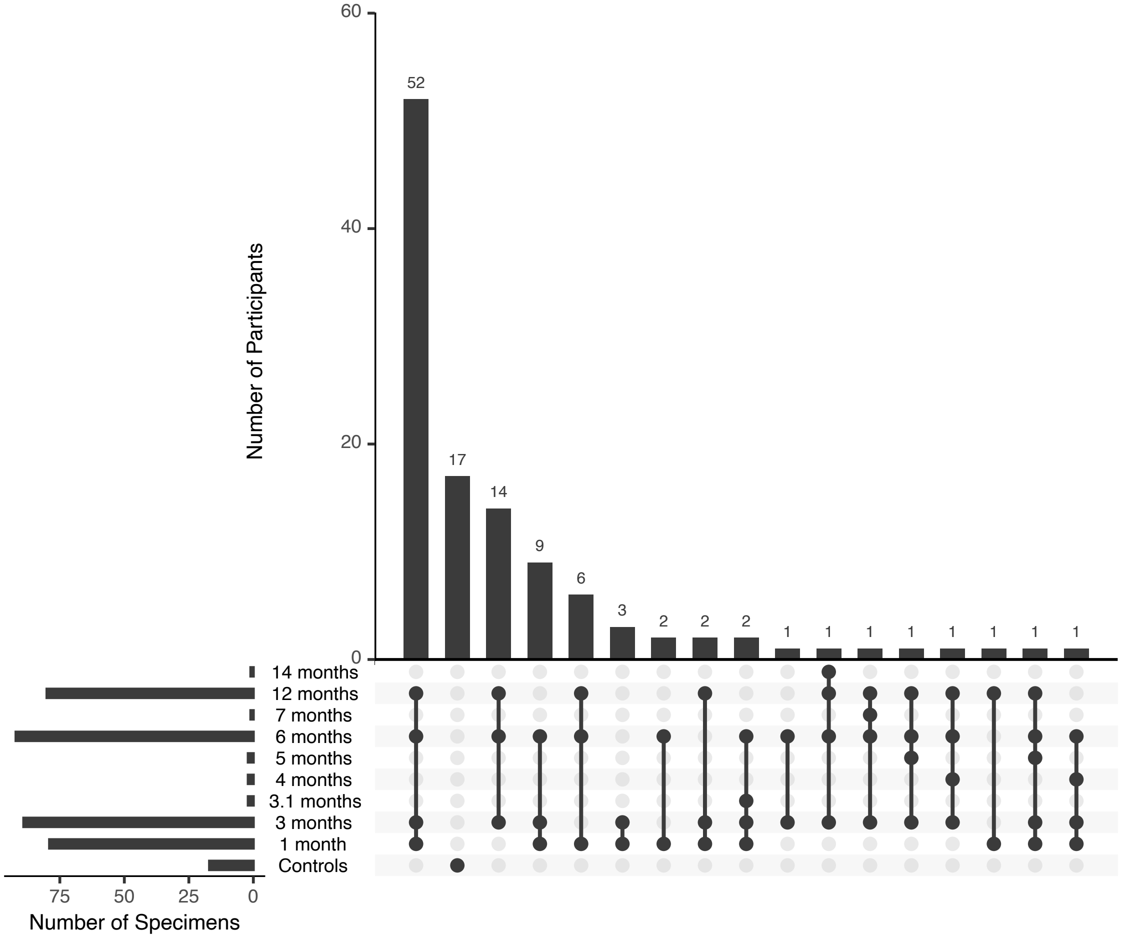


**Supplemental Figure 1.** Horizontal bar plots show the number of specimens collected for main timepoints: 1, 3, 6, and 12-months along with other peripheral timepoints. Vertical bar plots show the number of participants with a unique set of timepoints denoted by the connecting dots.

**Supp. Fig. 2**
