## Supplemental Figure 2 for "SARS-CoV-2 Humoral Immune Responses in Convalescent Individuals Over 12 Months Reveal Severity-Dependent Antibody Dynamics"

**Supp. Fig. 2**

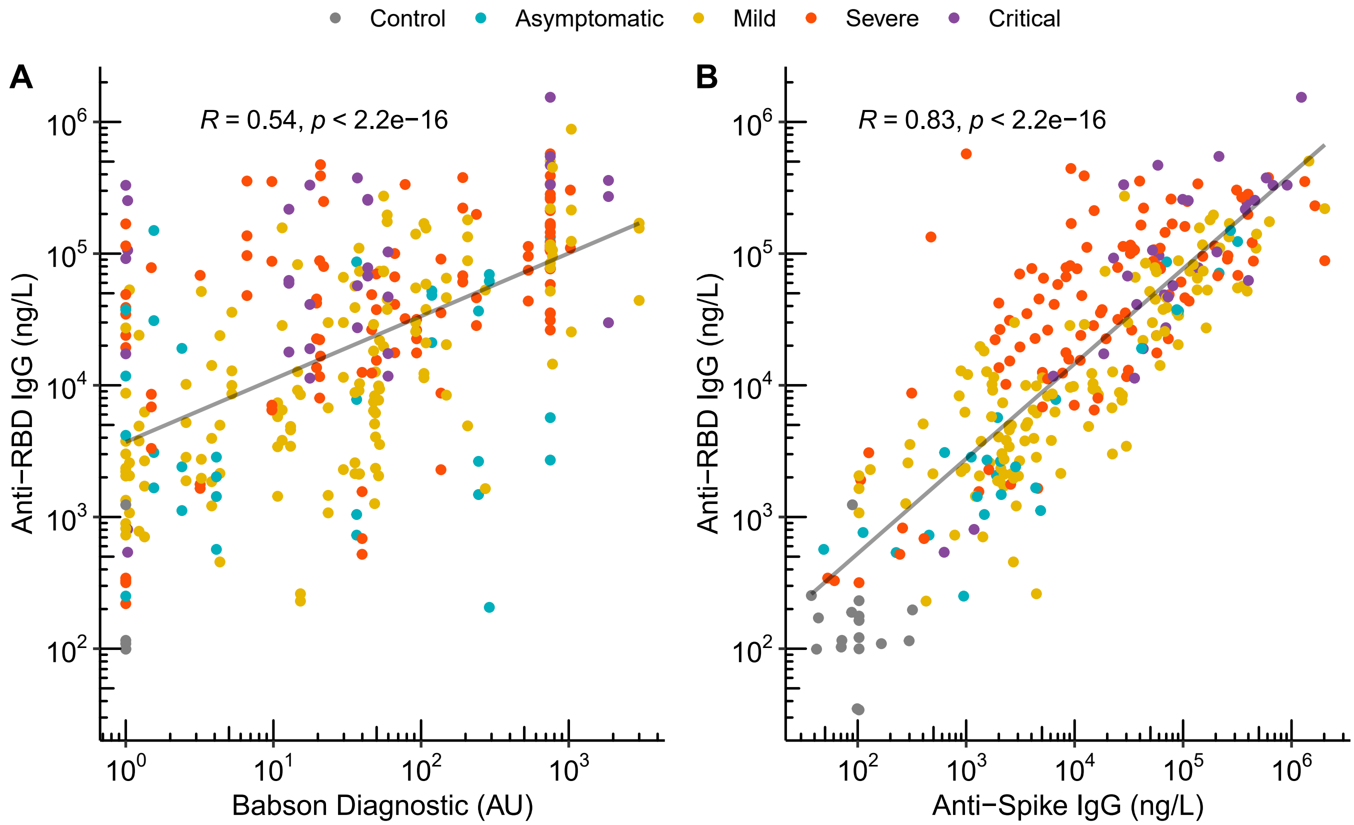


**Supplemental Figure 2.** Spearman correlation between anti-RBD IgG (BioLegend LEGEND MAX ELISA kit) and **A)** Babson Diagnostic (Siemens Healthineers Atellica IM sCOVG assay) **B)** anti-Spike IgG (lab-developed ELISA). Includes collections that proceeded a reinfection or vaccination.
